# Rapid identification of SARS-CoV-2-infected patients at the emergency department using routine testing

**DOI:** 10.1101/2020.04.20.20067512

**Authors:** Steef Kurstjens, Armando van der Horst, Robert Herpers, Mick W.L. Geerits, Yvette C.M. Kluiters-de Hingh, Eva-Leonne Göttgens, Martinus J.T. Blaauw, Marc H.M. Thelen, Marc G.L.M. Elisen, Ron Kusters

## Abstract

**Background:** The novel coronavirus disease 19 (COVID-19), caused by SARS-CoV-2, spreads rapidly across the world. The exponential increase in the number of cases has resulted in overcrowding of emergency departments (ED). Detection of SARS-CoV-2 is based on an RT-PCR of nasopharyngeal swab material. However, RT-PCR testing is time-consuming and many hospitals deal with a shortage of testing materials. Therefore, we aimed to develop an algorithm to rapidly evaluate an individual’s risk of SARS-CoV-2 infection at the ED.

**Methods:** In this multicenter retrospective study, routine laboratory parameters (C-reactive protein, lactate dehydrogenase, ferritin, absolute neutrophil and lymphocyte counts), demographic data and the chest X-ray/CT result from 967 patients entering the ED with respiratory symptoms were collected. Using these parameters, an easy-to-use point-based algorithm, called the corona-score, was developed to discriminate between patients that tested positive for SARS-CoV-2 by RT-PCR and those testing negative. Computational sampling was used to optimize the corona-score. Validation of the model was performed using data from 592 patients.

**Results:** The corona-score model yielded an area under the receiver operating characteristic curve of 0.91 in the validation population. Patients testing negative for SARS-CoV-2 showed a median corona-score of 3 versus 11 (scale 0-14) in patients testing positive for SARS-CoV-2 (p<0.001). Using cut-off values of 4 and 11 the model has a sensitivity and specificity of 96% and 95%, respectively.

**Conclusion:** The corona-score effectively predicts SARS-CoV-2 RT-PCR outcome based on routine parameters. This algorithm provides the means for medical professionals to rapidly evaluate SARS-CoV-2 infection status of patients presenting at the ED with respiratory symptoms.

## INTRODUCTION

In December 2019 the novel coronavirus disease 2019 (COVID-19), caused by SARS-CoV-2, spread rapidly from its origin in Wuhan, China (1). Symptoms can range from mild, common cold-like, to life threatening with intensive care unit admission and extensive mechanical ventilation (2, 3). On February 27^th^ 2020 the first patient was identified in the Netherlands, and thousands of new patients were diagnosed within the first month.

The subsequent exponential increase in prevalence has resulted in overcrowding of emergency departments (ED) and has led to a shortage of isolation rooms (4). For correct triaging of patients diagnostic testing is of key importance. The leading standard test for detecting SARS-CoV-2 is an RT-PCR of nasopharyngeal swab material (5). However, RT-PCR testing is time-consuming and shortage of testing materials and capacity imposes a serious threat (6).

Doctors at the ED are required to assess the probability of SARS-CoV-2 infection in each patient entering the ED. To accelerate the triage process at the ED, we integrated routine demographic, laboratory and imaging data of patients presenting at the ED with COVID-19-like symptoms to develop a point-based algorithm. This algorithm can assess whether a person, presenting at the ED with respiratory symptoms, is likely to have COVID-19. In case of a shortage of testing capacity, adoption of this algorithm could reduce the number of patients for whom RT-PCR testing is required. Moreover, implementation of the corona-score enables rapid decision making at the ED, lowering pressure on isolation rooms.

## METHODS

### Patient population

In this retrospective multicenter study, 375 patients from three different hospitals presenting at the ED with respiratory symptoms and subsequent SARS-CoV-2 RT-PCR testing were included (Figure 1 and Table 2). Patients from other departments and patients without any respiratory symptoms or suspicion of COVID-19 were excluded. An independent cohort of 592 patients from four hospitals was used to validate the model (Figure 1 and Table 2). For the validation population, patients with missing values or hemolytic samples were excluded (n=97).

**Table 1.** The point-based scoring system for the calculation of the corona-score. The final score is clamped from a minimum of 0 to a maximum of 14 points. More information can also be found at www.corona-score.com.

|  |  |  |  |  |  |  |  |
| --- | --- | --- | --- | --- | --- | --- | --- |
| Age (years) | ≤75 | 76-79 | 80+ |  |  |  |  |
| Points | 0 | 1 | 2 |  |  |  |  |
| Sex | Female | Male |  |  |  |  |  |
| Points | 0 | 1 |  |  |  |  |  |
| CRP (mg/L) | 0-9 | 10-14 | 15-38 | 39-69 | 70-193 | 194-303 | 304+ |
| Points | 0 | 1 | 2 | 3 | 2 | 1 | 0 |
| Ferritin (µg/L) | ≤15 | 16-179 | 180-301 | 302-538 | ≥539 |  |  |
| Points | -1 | 0 | 1 | 2 | 3 |  |  |
| LDH (U/L) | ≤257 | 258-265 | 266-397 | ≥398 |  |  |  |
| Points | 0 | 1 | 2 | 3 |  |  |  |
| ALC (10<sup>9</sup>/L) | ≤1.2 | ≥1.3 |  |  |  |  |  |
| Points | 1 | 0 |  |  |  |  |  |
| ANC (10<sup>9</sup>/L) | ≤5.1 | 5.2-7.9 | 8.0-9.0 | 9.1-10.3 | ≥10.4 |  |  |
| Points | 0 | -1 | -2 | -3 | -4 |  |  |
| CXR | No infiltrate |  | Unilateral infiltrate |  | Bilateral infiltrate |  |  |
| Points | 0 |  | 1 |  | 4 |  |  |

CRP, C-reactive protein; LDH, lactate dehydrogenase; ALC, absolute lymphocyte count; ANC, absolute neutrophil count; CXR, chest X-ray.

**Table 2.**
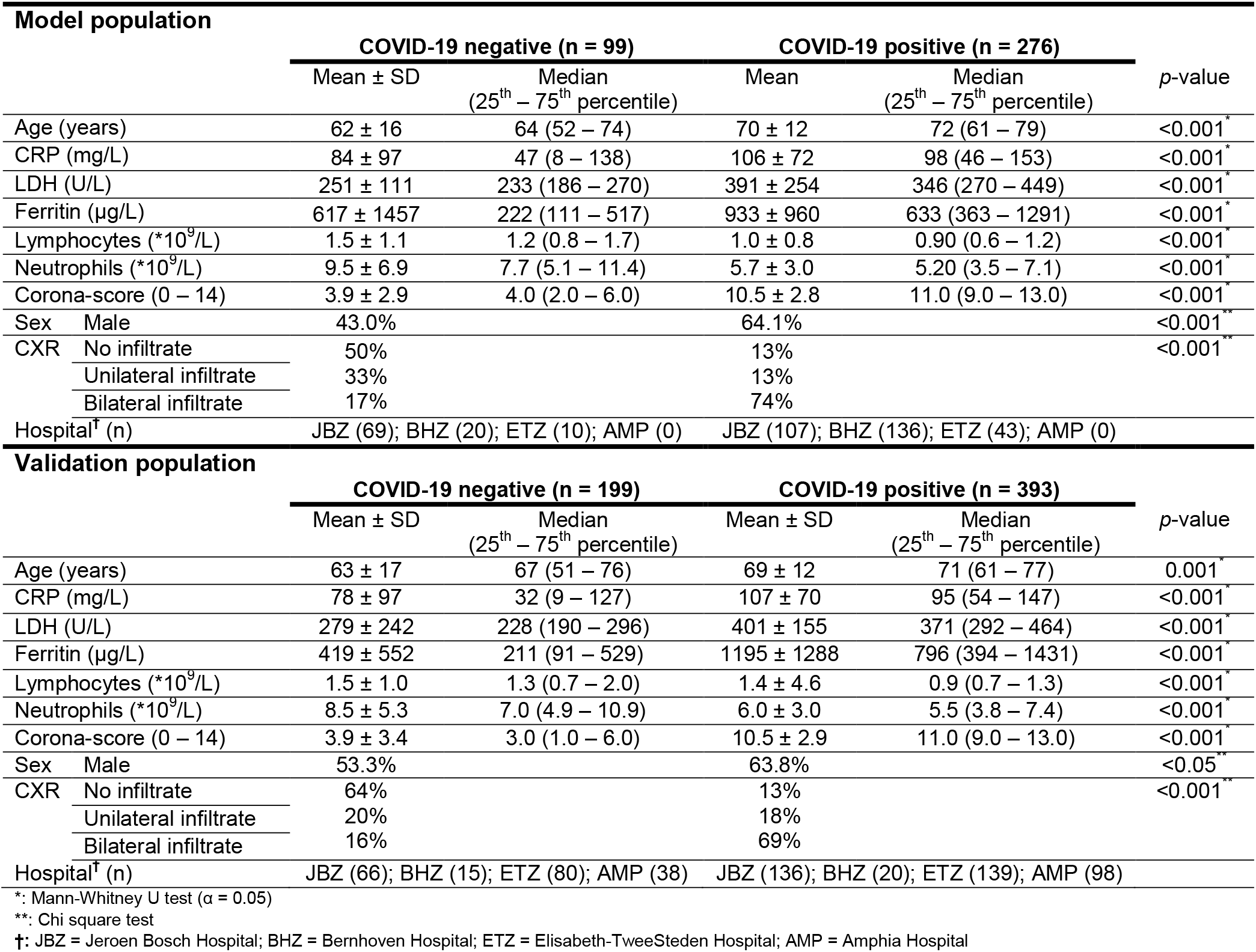
Overview of the demographic, clinical chemistry and chest x-ray parameters of the patients included in the model development and validation.

**Figure 1.**
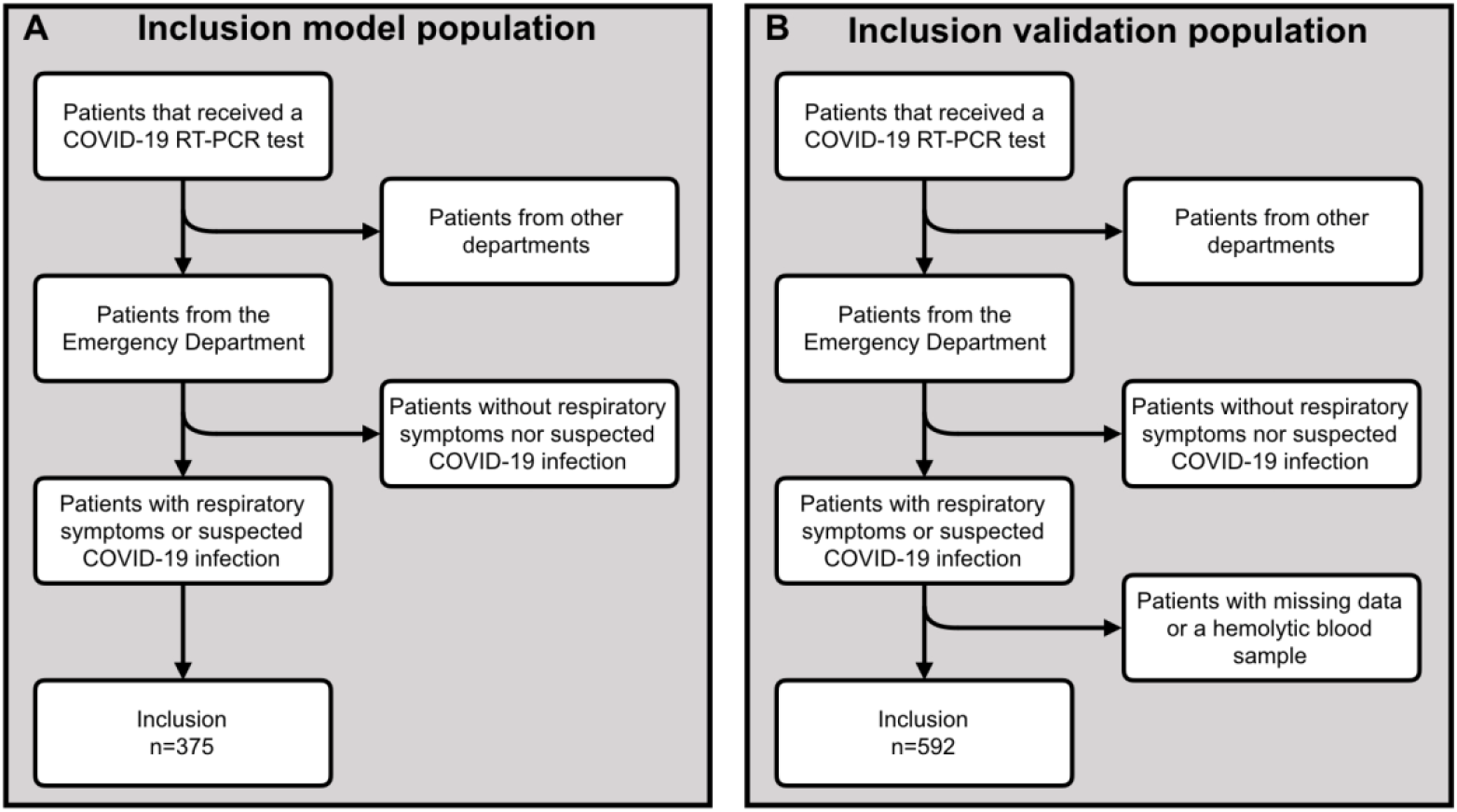
Flow diagram of the study setup for the model and validation population. (A) A flow diagram depicting the inclusion and exclusion of patients that were used to develop the algorithm. A total of 375 patients were included. (B) A flow diagram depicting the inclusion and exclusion of patients that were used to validate the algorithm. A total of 592 patients were included.

### Measurements

For clinical chemistry analyses and RT-PCR, venous blood and pharyngeal plus nasal swab specimens, respectively, were collected. Clinical chemistry parameters (C-reactive protein (CRP), ferritin, lactate dehydrogenase (LDH), absolute lymphocyte and neutrophil counts (ALC and ANC)) were obtained on routine analyzers from Siemens (Jeroen Bosch Hospital and the (immuno-)chemistry of Bernhoven Hospital), Sysmex (Elisabeth TweeSteden Hospital and the hematology of Amphia Hospital), Roche (Elisabeth TweeSteden Hospital and the (immuno-)chemistry of Amphia Hospital) and Abbott (hematology of Bernhoven Hospital). SARS-CoV-2 RT-PCR testing at Amphia Hospital and Elizabeth TweeSteden Hospital was performed using tests from Microvida Laboratory (the Netherlands), whereas Jeroen Bosch Hospital and Bernhoven Hospital used in-house developed tests. Chest X-rays (CXR) and chest CT-scans were imaged using Siemens, GE Healthcare and Philips equipment. External quality assessment (EQA) in commutable materials by Dutch Foundation for Quality Assessment in Medical Laboratories (SKML) demonstrated that ferritin measured on Roche analyzers is on average 20% higher than on Siemens analyzers. For building the model and calculating corona-scores ferritins measured on Siemens analyzers were therefore multiplied by 1.2. All other measurands in the scoring system had no significant inter-method differences for Roche, Siemens and Sysmex in the particular SKML EQA schemes.

### Corona-score algorithm

A scoring-based algorithm was developed using laboratory measurands (CRP, ALC, ANC, LDH and ferritin), age, sex and CXR/CT as input. Scores were assigned to each parameter according to Table 1 (or see www.corona-score.nvkc.nl) for more information). The corona-score is obtained by the summation of the score for each parameter. The final score is clamped from a minimum of 0 to a maximum of 14 points. Cut-off points and weights of demographic, laboratory and imaging parameters were computationally sampled using Python (v3.7.0, Python Software Foundation, USA) to optimize for a maximum area under the receiver operating characteristic (AUROC) curve. When values were missing in the data of the model population (n=3 for ALC and ANC, n=31 for LDH and n=4 for ferritin) the median of the total population was used.

### Statistical analyses

Data were analyzed using the Excel 2010 (Microsoft Corporation, USA) plugin ‘Analyse-it v5.11’ (Analyse-it Software, Ltd, UK) and SPSS statistics v22 (IBM, USA). Continuous variables were tested for normal distribution using a Kolmogorov-Smirnov test. In case of non-normal distribution, a Mann-Whitney U test was performed to compare the medians. Categorical variables were compared by a chi square test. A *p*-value <0.05 was considered statistically significant.

## RESULTS

Using a cohort of 375 ED patients with respiratory symptoms a point-based algorithm was created and subsequently validated using a separate independent cohort of 592 patients (Table 2). At the time of presentation at the ED the parameters sex, age, CRP, ferritin, LDH, ALC, ANC and CXR were significantly different between the COVID-19 positive and negative patients (Figure 2 and Table 2). Together, these parameters were used to develop an algorithm, named ‘corona-score’. Inclusion of albumin, procalcitonin or clinical parameters such as fever, cough and dyspnea did not sufficiently improve the performance of the algorithm (data not shown). The corona-score resulted in a model with an AUROC of 0.94 (Figure 3A, 95% CI 0.91 – 0.96). Patients with a negative RT-PCR test had a median of 4 compared to a median of 11 for SARS-CoV-2 positive patients (Figure 3B and Table 2). The corona-score algorithm was validated with data from 592 patients, yielding an AUROC of 0.91 (Figure 3C, 95% CI 0.89-0.94). In the validation population SARS-CoV-2 negative patients had a median of 3 versus a median of 11 for SARS-CoV-2 positive patients (Figure 3D and Table 2).

**Figure 2.**
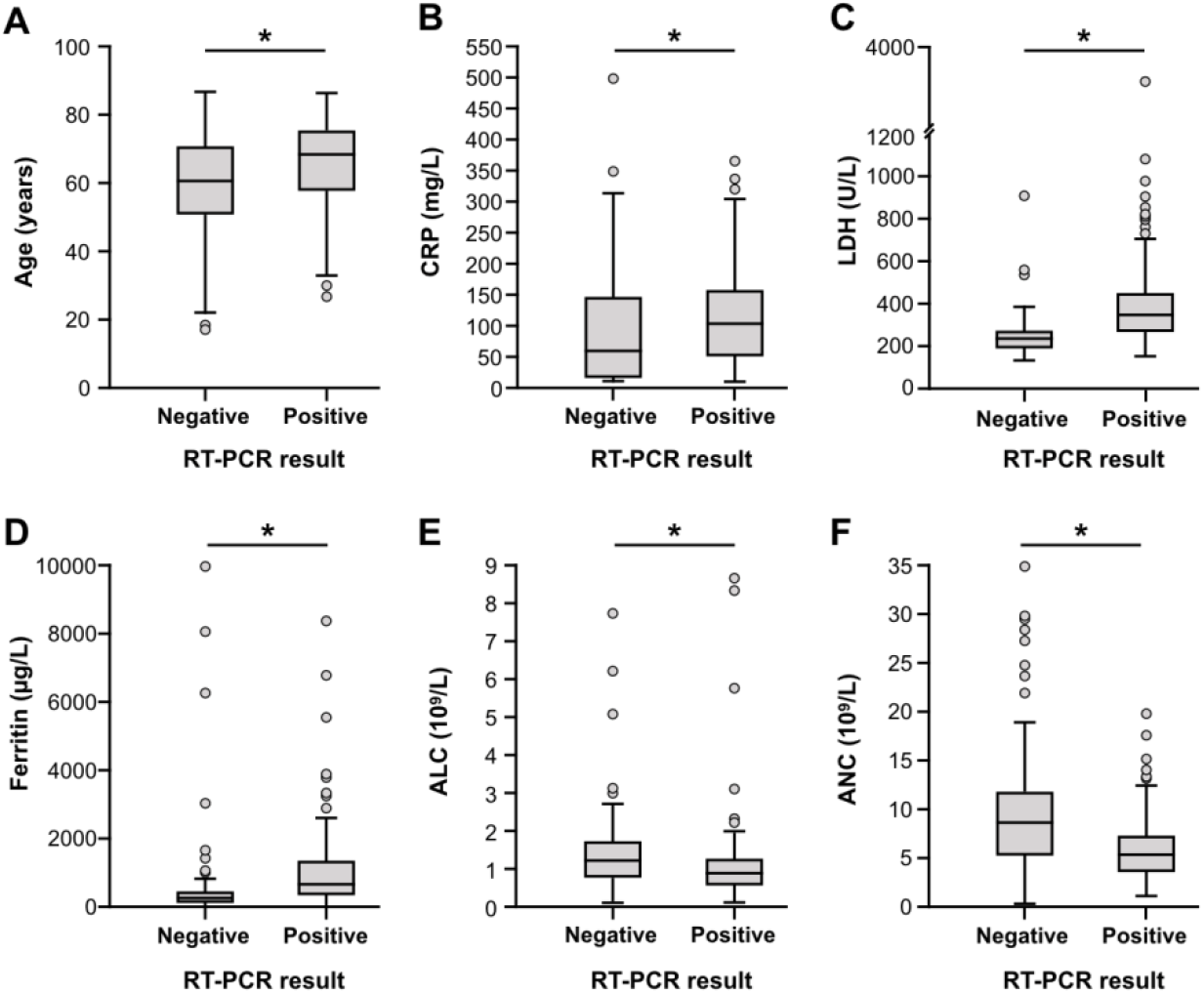
Difference in demographic and routine laboratory parameters between COVID-19 positive and negative patients. Box plots depicting the median and interquartile range of continuous variables included in our model, (A) age, (B) C-reactive protein (CRP), (C) lactate dehydrogenase (LDH), (D) ferritin, (E) absolute lymphocyte count (ALC), (F) absolute neutrophil count (ANC). Data are taken from the model population presented in Table 2. * indicates a *p*-value ≤0.05.

**Figure 3.**
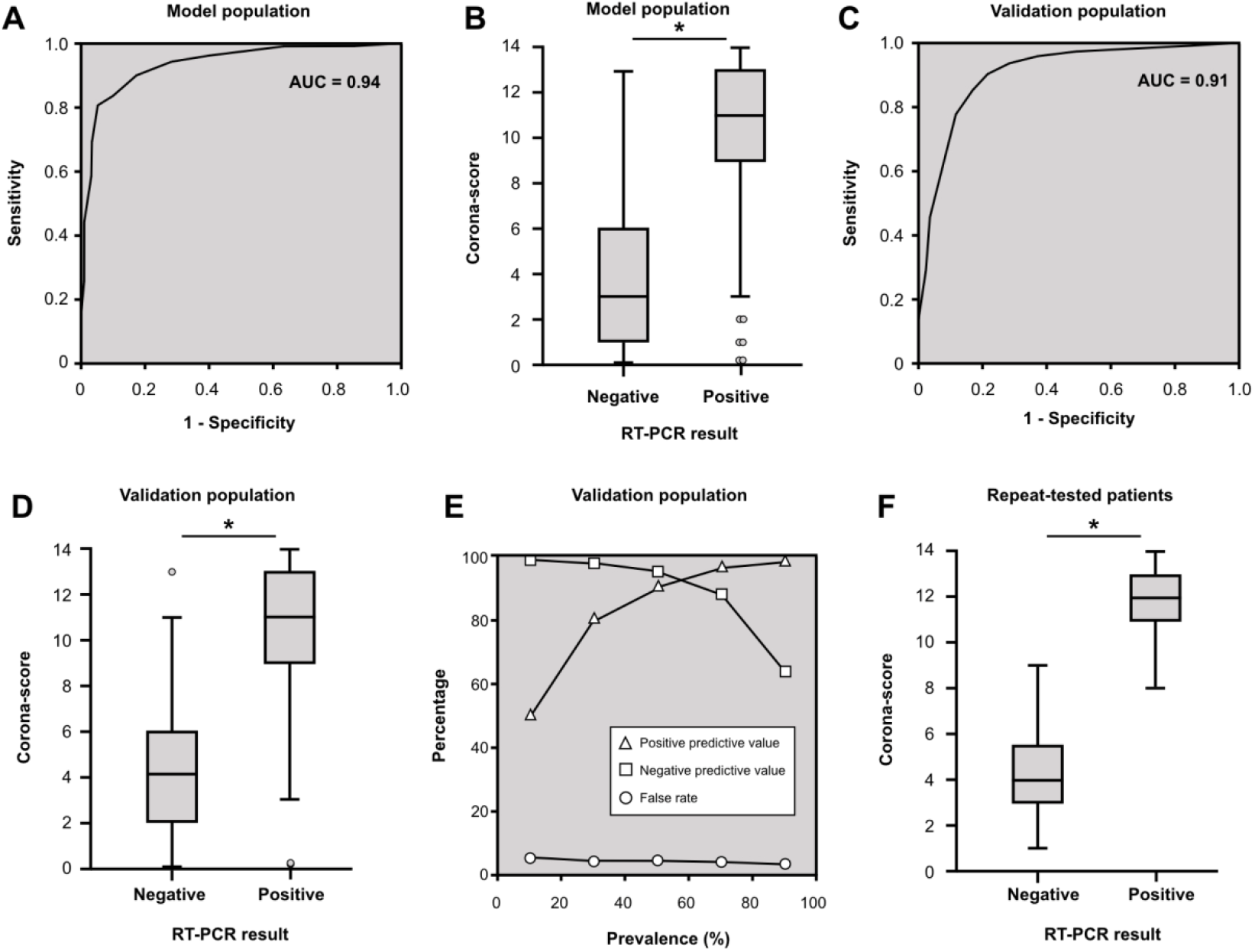
Performance of the corona-score to predict RT-PCR outcome. (A) ROC-curve (AUROC = 0.94, 95% CI 0.91 – 0.96) of the model, created using data from 375 patients from 3 different hospitals. Points were attributed to each patient based on demographic, laboratory and CXR data (the range of the corona-score is clamped from 0 – 14). (B) Box-plot displaying the difference in the median between the SARS-CoV-2 negative and positive patients from the model population obtained using the corona-score. (C) The model was validated using 592 patients (AUROC = 0.91, 95% CI 0.89-0.94). (D) Box-plot displaying the difference in the median between the SARS-CoV-2 negative and positive patients from the validation population obtained using the corona-score. (E) Positive (triangle) and negative (square) predictive values and false rate (circle) at several different prevalences, using a corona-score of four and eleven as lower and upper cut-offs, respectively. (F) Box-plot depicting the median corona-score of patients that received multiple SARS-CoV-2 RT-PCR tests, for whom the latest RT-PCR (material obtained from either nasopharyngeal, fecal or sputum) was positive (n=13) or remained negative (n=12). * indicates a *p*-value ≤0.05.

By using different cut-off values the desired sensitivity and specificity for the test can be found (Table 3). Using corona-score cut-offs of 4 (96% sensitivity) and 11 (95% specificity) at a 70% prevalence, this model showed negative and positive predictive values of 88% and 96% (Figure 3E). The total false rate given these conditions is 4% (Figure 3E).

**Table 3.**
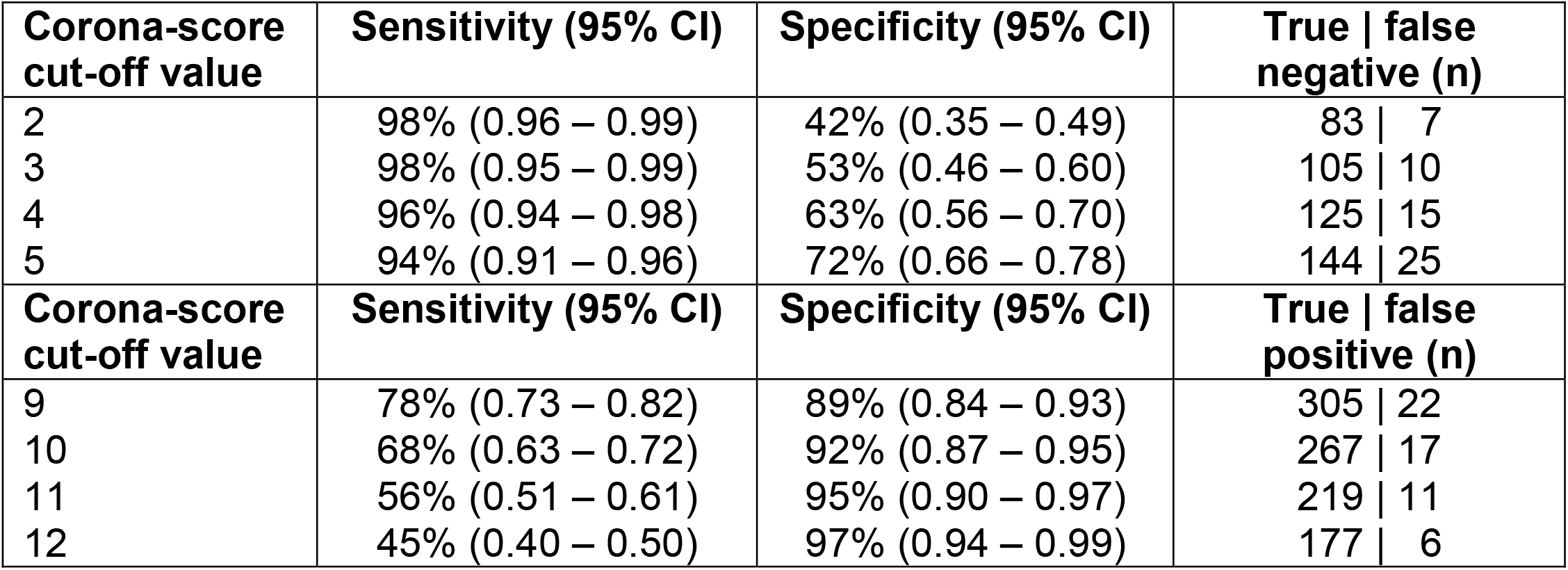
Sensitivity and specificity at different lower and upper cut-off values for the corona-score (value included, ≤ for 2 to 5 and ≥ for 9 to 12) determined using the validation population (n = 592). The right column depicts the number of true and false negative and positive patients.

RT-PCR testing for SARS-CoV-2 is hampered by significant numbers of false negatives as the sensitivity of RT-PCR is estimated at approximately 70-90% (7). Indeed, many doctors request multiple COVID-19 tests when the RT-PCR result does not match the clinical presentation of the patient. Patients from the validation population of the JBZ hospital that showed positivity for SARS-CoV-2 after repeated RT-PCR testing (n=13) had an initial median corona-score of 12, while patients that remained negative (n=12) had an initial median corona-score of 4 (Figure 3F). This shows that the corona-score is able to distinguish between true and false negatives.

## DISCUSSION

Using a cohort of 967 patients we developed and validated a point-based algorithm to predict the likelihood of SARS-CoV-2 infection in patients presenting at the ED with respiratory symptoms. Validation of the model resulted in an AUROC of 0.91 with a 96% sensitivity and 95% specificity, using corona-score cut-offs of 4 and 11. Such an algorithm can be used to, 1) accelerate determination of isolation needs and 2) reduce RT-PCR testing: a reduction of about 60% can be achieved if cut-offs of 4 and 11, yielding 125 true negative and 219 true positive patients in the validation cohort of 592 patients, are used.

Our algorithm is optimized to predict the outcome of the SARS-CoV-2 RT-PCR test, which has limited (70-90%) sensitivity (7). Inclusion of patients having false negative RT-PCR tests into the validation population results in an underestimation of the performance of the algorithm. Interestingly, for twenty-five patients that received multiple COVID-19 tests our algorithm could predict which patients were initially false negatives. Therefore, the sensitivity of the corona-score appears to exceed the sensitivity of the initial SARS-CoV-2 RT-PCR.

In a minority of cases, our model produces a corona-score of 0 – 5 in patients that tested positive for SARS-CoV-2 by RT-PCR. There are two common underlying reasons for this phenomenon. Firstly, the corona-score performed poorly in patients with a gastro-intestinal presentation of COVID-19, but without respiratory symptoms. Therefore, this algorithm should only be used for patients at the ED with respiratory symptoms. Secondly, patients that only have mild respiratory symptoms, and therefore do not have large alterations in their laboratory parameters, generally have a low corona-score. However, in most cases the patients with a mild presentation were not hospitalized. Therefore, we consider that the low corona-score corresponds with the clinical findings. On the other hand, some negatively-tested patients received a high corona-score. This could be due to false-negative RT-PCR testing or possibly other viral infections. Interestingly, four patients that were positive for influenza and negative for SARS-CoV-2 had a low corona-score (2 – 6). During this COVID-19 pandemic, the prevalence of other respiratory viruses appears very low; hence, the discriminative potential of the corona-score in patients infected by such viruses could not be systematically established. Notably, in case of any viral outbreak, a similar modelling approach could be considered to develop an algorithm as described here.

The four laboratories involved in this study deploy different instruments from the major *in-vitro* diagnostic device providers. Most measurands that were included in the algorithm have an identical metrological traceability and hence comparable results in the commutable EQA scheme of the SKML (8). However, there is no reference method for ferritin (9). The different calibrations lead to approximately 20% difference in ferritin results between the methods employed by the laboratories in this study. Therefore, a 1.2 harmonization factor was applied to the ferritin values obtained from Siemens instruments, before calculating corona-scores, correcting the lack of standardization. Generally, methodological harmonization between laboratories should be encouraged for better comparison of laboratory results (10).

To our knowledge, our algorithm is the first available validated tool to rapidly evaluate COVID-19 status in ED patients with respiratory symptoms based on routine laboratory tests. The model has already been implemented at the ED of several hospitals in the Netherlands. Implementation of this algorithm will accelerate the triage of patients and reduce the number of RT-PCR tests required.

## Data Availability

Data availability
The data that support the findings of this study are available from the corresponding author upon reasonable request. More information can be obtained at www.corona-score.nvkc.nl.

## Author Contributions

All authors confirmed they have contributed to the intellectual content of this paper and have met the following 4 requirements: (a) significant contributions to the conception and design, acquisition of data, or analysis and interpretation of data; (b) drafting or revising the article for intellectual content; (c) final approval of the published article; and (d) agreement to be accountable for all aspects of the article thus ensuring that questions related to the accuracy or integrity of any part of the article are appropriately investigated and resolved.

## Acknowledgments

We thank Rob Verheyen (University College London), Thomas de Bel (Radboudumc Nijmegen), Marleen Weijland, Daniëlle Verboogen PhD (Elizabeth TweeSteden Hospital), Kinki Jim MD (Jeroen Bosch Hospital), dr. Rein Hoedemakers (Jeroen Bosch Hospital) and Jasmijn van Balveren MD (Jeroen Bosch Hospital) for their valuable technical help and scientific input.

## Statement of ethics

The study was conducted according to the declaration of Helsinki, Guidelines for Good Clinical Practice and the Dutch Medical Research Involving Human Subjects Act. The execution of this retrospective observational study of patient records was approved by the local review board of the Jeroen Bosch Hospital. This study had no effect on the behaviour of patients or medical decision-making.

## Data availability

The data that support the findings of this study are available from the corresponding author upon reasonable request. More information can be obtained at www.corona-score.nvkc.nl.

## List of Abbreviations

ALC: Absolute lymphocyte count
AMP: Amphia Hospital
ANC: Absolute neutrophil count
AUROC: Area under the receiver operating characteristic
BHZ: Bernhoven Hospital
COVID-19: Coronavirus disease 19
CRP: C-reactive protein
CXR: Chest X-ray
ED: Emergency department
EQA: External quality assessment
ETZ: Elizabeth TweeSteden Hospital
JBZ: Jeroen Bosch Hospital
LDH: Lactate dehydrogenase
SKML: Dutch Foundation for Quality Assessment in Medical Laboratories

